# Antibiotic prescriptions in children with COVID-19 and Multisystem Inflammatory Syndrome: a multinational experience in 990 cases from Latin America

**DOI:** 10.1101/2020.12.05.20243568

**Authors:** Adriana Yock-Corrales, Jacopo Lenzi, Rolando Ulloa-Gutiérrez, Jessica Gómez-Vargas, Antúnez-Montes Omar Yassef, Jorge Alberto Rios Aida, Olguita del Aguila, Erick Arteaga-Menchaca, Francisco Campos, Fadia Uribe, Andrea Parra Buitrago, Lina Maria Betancur Londoño, Martin Brizuela, Danilo Buonsenso

**Affiliations:** Pediatric Emergency Department, Hospital Nacional de Niños “Dr. Carlos Sáenz Herrera”, CCSS, San José, Costa Rica; Department of Biomedical and Neuromotor Sciences, Alma Mater Studiorum - University of Bologna, Bologna, Italy; Infectious Disease Department. Hospital Nacional de Niños “Dr. Carlos Sáenz Herrera”, CCSS, San José, Costa Rica; Departamento de Docencia e Investigación, Instituto Latinoamericano de Ecografía en Medicina (ILEM)., Ciudad de Mexico, Mexico; CLÍNICA JAS MÉDICA, Lima, Perù; Unidad de Infectología Pediátrica del Hospital Nacional Edgardo Rebagliati Martins-Lima-Perú; Hospital General Regional 200 IMSS, Mexico; Hospital Madre Niño San Bartolome, Lima, Peru; Hospital Pablo Tobon Uribe Medellin, Colombia; Fundacion Neumologica Colombiana,Bogotà, Colombia; Pediatric Infectious Disease, Hospital isidoro Iriarte, Quilmes, Buenos Aires, Argentina; Department of Woman and Child Health and Public Health, Fondazione Policlinico Universitario A. Gemelli, Rome, Italy; Dipartimento di Scienze Biotecnologiche di Base, Cliniche Intensivologiche e Perioperatorie, Università Cattolica del Sacro Cuore, Rome, Italy; Global Health Research Institute, Istituto di Igiene, Università Cattolica del Sacro Cuore, Roma, Italia

**Author notes:** Email addresses: Adriana Yock-Corrales, Jacopo Lenzi, Rolando Ulloa-Gutiérrez, Jessica Gómez-Vargas, Antúnez-Montes Omar Yassef, Jorge Alberto Rios Aida, Olguita del Aguila, Erick Arteaga-Menchaca, Francisco Campos, Fadia Uribe, Andrea Parra Buitrago, Lina Maria Betancur Londoño, Martin Brizuela, Danilo Buonsenso.

**Keywords:** COVID-19, SARS-CoV-2, MIS-C, antibiotic stewardship

## Abstract

**Background:** To date, there are no comprehensive data on antibiotic use in children with COVID-19 and Multisystem Inflammatory Syndrome (MIS-C).

**Methods:** Multicenter cohort study from 5 Latin American countries. Children 17 years of age or younger with microbiologically confirmed SARS-CoV-2 infection or fulfilling MIS-C definition were included. Antibiotic prescriptions were collected and factors associated with their use were calculated.

**Findings:** 990 children were included, with a median age of 3 years (interquartile range 1–9). Of these, 69 (7.0%) were diagnosed with MIS-C. The prevalence of antibiotic use was 24.5% (*n* = 243). MIS-C with (OR = 45.48) or without (OR = 10.35) cardiac involvement, provision of intensive care (OR = 9.60), need for hospital care (OR = 6.87), pneumonia and/or ARDS detected through chest X-rays (OR = 4.40), administration of systemic corticosteroids (OR = 4.39), oxygen support, mechanical ventilation or CPAP (OR = 2.21), pyrexia (OR = 1.84), and female sex (OR = 1.50) were independently associated with increased use of antibiotics. On the contrary, lower respiratory tract infections without radiologic evidence of pneumonia/ARDS and not requiring respiratory support (OR = 0.34) were independently associated with decreased use of antibiotics. There was significant variation in antibiotic use across the hospitals.

**Conclusions:** Our study showed a relatively high rate of antibiotic prescriptions in children with COVID-19 and in particular in those with severe disease or MIS-C. Importantly, we found a significant variation in reasons for prescriptions of antibiotics and type of chosen therapies, as well in hospital practices, highlighting current uncertainties and lack of guidelines for the recognition of bacterial infections in children with COVID-19. Prospective studies are needed to provide better evidence on the recognition and management of bacterial infections in COVID-19 children.

**What is known:** COVID-19 may worsen antibiotic prescription practices

**What this new:** COVID-19 and MIS-C children frequently received antibiotics

There was a wide variation in antibiotic prescriptions among institutions, highlighting the lack of practicle guidelines in the use of antibiotics in children with COVID-19

## Introduction

Months after the first description of COVID-19 in China, growing evidence is raising about the impact of SARS-CoV-2 infection on the pediatric population. Several studies from China (1), Europe (2,3), United States (4) and Latin America (5) are clarifying that COVID-19 in children is typically mild, although patients with medically complex conditions or those of minority race/ethnicity deserve more attention because they may be at risk of more severe disease (4). The Multisystem Inflammatory Syndrome (MIS-C), an entity not yet fully clarified related to SARS-CoV-2, is a severe complication of the exposition to the virus, which may require Intensive Care Admission, mechanical ventilation and cardio-respiratory support, rarely leading to death (6). Because SARS-CoV-2 is a viral infection, and the resulting disease is usually mild in children, it is not expected that a child with COVID-19 would routinely receive antimicrobials. This is particularly true for the second period of the pandemic, when the non-utility of azithromycin, initially suggested as a drug with potential anti-viral properties (7), has been showed (8). The MIS-C can be an exception to this concept, since the severe and acute presentation may be similar to the toxic-shock syndrome and available consensus documents suggest empiric wide-spectrum antibiotic therapy until bacterial infections are ruled-out (9).

Nevertheless, there are growing concerns about the possible negative impact of the pandemic on antimicrobial use. While this is particularly discussed for adults with COVID-19 (10), Velasco-Arnaiz et al (11) reported preliminary data suggesting that the pandemic has the potential to have a significant impact on antimicrobial use in the pediatric inpatient population. They did assess antibiotic prescriptions during and before the pandemic, but did not assess directly antibiotic use and its determinants in COVID-19 children.

Since cases are constantly raising worldwide, it is expected that SARS-CoV-2 will circulate still for a long time, therefore the appropriate management of children with COVID-19 is a priority. While the pandemic only determined a limited direct impact on children, inappropriate prescriptions have the potential of worsening an already dangerous situation, i.e. antimicrobial resistance. Due to the gap in available literature, we performed a multinational study in Latin America aiming to assess the use of antibiotics in children with COVID-19 and understand the determinants of its use.

## MATERIALS AND METHODS

### Study Design and Participants

This study is part of an ongoing independent project assessing COVID-19 and MIS-C in Latin American children, already presented elsewhere (12) and with a previous published paper describing an initial group of 409 children with confirmed COVID-19 (5). For the current study, we aimed to assess determinants of antibiotic use in children with COVID-19. We implemented the previously used dataset (2, 5) including data regarding name of antibiotic used and the reason why the attending clinician decided to administer antibiotics. The remaining variables are those previously used and included age, gender, symptoms, imaging, underlying medical conditions, need for hospital and NICU/PICU admission, respiratory and cardiovascular support, other viral co-infections, drugs used to treat COVID-19, development of MIS-C and type of organ involvement, and outcome.

The study was reviewed and approved by the CoviD in sOuth aMerIcaN children—study GrOup core group and approved by the Ethics Committee of the coordinating center and by each participating center (Mexico: COMINVETICA-30072020-CEI0100120160207; Colombia: PE-CEI-FT-06; Peru: No. 42-IETSI-ESSALUD-2020; Costa Rica: CEC-HNN-243-2020). The study was conducted in accordance with the Declaration of Helsinki and its amendments. No personal or identifiable data were collected during the conduct of this study.

### Statistical analysis

Summary statistics for the study sample were presented as counts and percentages. The association of relevant demographic and clinical characteristics with antibiotic use was assessed using a multivariable logistic regression model; the effect size of covariates was expressed by odds ratios (ORs) with 95% confidence intervals (CIs). The variables considered in this analysis were age, sex, medical history of immunodeficiency, immunosuppressants or chemotherapy, hospital care, pyrexia, upper and lower respiratory tract infections, gastrointestinal symptoms, headache, chest X-ray abnormalities, respiratory support, administration of systemic corticosteroids, and diagnosis of MIS-C, both with and without cardiac involvement. A set of dummy variables for individual hospitals was also included in the model to adjust for the potential bias of confounding by center. All data were analyzed using the Stata 15 software (StataCorp. 2017. *Stata Statistical Software: Release 15*. College Station, TX: StataCorp LLC). The significance level was set at 5% and all tests were 2-sided.

### Dataset availability

The dataset generated for this study is available upon request to the corresponding author.

## RESULTS

### Study population

A total of 921 children (93.0%) with microbiologically confirmed COVID-19 and 69 children (7.0%) with MIS-C from Peru (*n* = 383, 38.7%), Costa Rica (*n* = 299, 30.2%), Argentina (*n* = 253, 25.6%), Colombia (*n* = 43, 4.3%) and Mexico (*n* = 12, 1.2%) were included in the study. The demographic and clinical characteristics of the 990 study patients are summarized in Table 1. The median age was 3 years (interquartile range: 1–9), ranging from 2 days to 17 years; 484 (48.9%) were female. The most common known source of transmission of the infection was a parent, considered the index case in 281 (28.4%) cases. A total of 303 (30.6%) children were admitted to hospital and 47 (4.7%) required admission to a Pediatric Intensive Care Unit (PICU). Fever was reported in 677 cases (68.4%); 466 (47.1%) children had symptoms suggestive of upper respiratory tract infection while 215 (21.7%) had lower respiratory tract symptoms; 301 (30.4%) had gastrointestinal symptoms. A chest radiograph was done in 285 (28.8%) patients. Of these, 92 (32.3%) had signs of COVID-19 pneumonia. Respiratory co-infections were detected in 14 (1.4%) children.

**Table 1.** Characteristics of the study sample (*n* = 990).

| Characteristic | *n* | % |
| --- | --- | --- |
| Female sex | 484 | 48.9 |
| Age group |  |  |
| 0 y | 202 | 20.4 |
| 1–2 y | 229 | 23.1 |
| 3–5 y | 144 | 14.5 |
| 6–11 y | 247 | 24.9 |
| 12–17 y | 168 | 17.0 |
| COVID-19 confirmed by real-time PCR | 639 | 64.5 |
| Positive SARS-CoV-2 IgG | 352 | 35.6 |
| Delay between onset and diagnosis |  |  |
| 0–1 d | 437 | 44.1 |
| 2–7 d | 460 | 46.5 |
| >7 d | 93 | 9.4 |
| Likely index case |  |  |
| Parent | 281 | 28.4 |
| Sibling | 14 | 1.4 |
| Other | 120 | 12.1 |
| Unknown | 575 | 58.1 |
| Medical history |  |  |
| Known history of BCG vaccine | 740 | 74.7 |
| Pre-existing medical conditions | 128 | 12.9 |
| Immunosuppressants at the time of diagnosis | 11 | 1.1 |
| Primary or secondary immunodeficiency | 8 | 0.8 |
| Chemotherapy over the last 6 months | 8 | 0.8 |
| Admitted to the hospital | 303 | 30.6 |
| Intensive care during hospital stay | 47 | 4.7 |
| Symptoms |  |  |
| Pyrexia ($\geq 38.0/\geq 100.4$ °C/°F) | 677 | 68.4 |
| Upper respiratory tract infection | 466 | 47.1 |
| Diarrhea and/or vomiting | 301 | 30.4 |
| Lower respiratory tract infection | 215 | 21.7 |
| Headache | 104 | 10.5 |
| Chest X-ray |  |  |
| Not performed | 705 | 71.2 |
| Negative | 193 | 19.5 |
| Positive (pneumonia* and/or ARDS†) | 92 | 9.3 |
| Respiratory support |  |  |
| Oxygen support | 117 | 11.8 |
| Mechanical ventilation | 31 | 3.1 |
| Continuous positive airway pressure (CPAP) | 11 | 1.1 |
| Extracorporeal membrane oxygenation (ECMO) | 0 | 0.0 |
| Administration of inotropes | 29 | 2.9 |
| Co-infections detected in respiratory samples(s)‡ | 14 | 1.4 |
| Drug administration |  |  |
| Systemic corticosteroids | 90 | 9.1 |
| Intravenous immunoglobulin (IVIG) | 60 | 6.1 |
| Hydroxychloroquine | 9 | 0.9 |
| Oseltamivir | 8 | 0.8 |
| Lopinavir or ritonavir | 3 | 0.3 |
| Non-corticosteroid immunosuppressants | 3 | 0.3 |
| Favipiravir | 2 | 0.2 |
| Remdesivir | 2 | 0.2 |
| Chloroquine, ribavirin or zanamivir | 0 | 0.0 |
| MIS-C diagnosis |  |  |
| No | 921 | 93.0 |
| Yes, with no cardiac or joint involvement | 33 | 3.3 |
| Yes, with cardiac involvement§ | 23 | 2.3 |
| Yes, with joint involvement | 11 | 1.1 |
| Yes, with cardiac and joint involvement§ | 2 | 0.2 |
| Tocilizumab administration to treat MIS-C | 8 | 0.8 |
| Current status |  |  |
| All symptoms resolved | 969 | 97.9 |
| Dead¶ | 8 | 0.8 |
| Still symptomatic | 7 | 0.7 |
| Long-term sequelae | 6 | 0.6 |
| Center |  |  |
| Peru | 383 | 38.7 |
| Costa Rica | 299 | 30.2 |
| Argentina | 253 | 25.6 |
| Colombia | 43 | 4.3 |
| Mexico | 12 | 1.2 |

\*45 cases of interstitial disease, 30 cases of consolidation, 4 cases of pleural effusion and 13 unspecified diagnoses.
‡3 cases of interstitial disease, 3 cases of consolidation and 10 unspecified diagnoses.
‡8 mycoplasmas, 3 rhinoviruses, 1 cytomegalovirus, 1 Epstein–Barr virus and 1 unspecified virus.
§10 cases of pericardial effusion, 6 cases of coronary dilatation, 5 cases of myocarditis and 4 cases of “other” cardiac involvement.
¶Mean time from symptom onset to death was $14 \pm 8$ days, ranging from 3 to 27.
COVID-19, coronavirus disease 2019; PCR, polymerase chain reaction; SARS-CoV-2, Severe acute respiratory syndrome coronavirus 2; IgG, immunoglobulin G; BCG, bacillus Calmette–Guérin; ARDS, Acute respiratory distress syndrome; MIS-C, multisystem inflammatory syndrome.

We found that 118 (11.9%) individuals required respiratory support; 117 (11.8%) required low-flow oxygen support, 11 (1.1%) were started on continuous positive airway pressure and 31 (3.1%) on mechanical ventilation. A total of 29 (2.9%) patients required inotropic support. Eight children died (0.8%). Further details described in table 1.

### Antibiotic use in COVID-19 and MIS-C children

The prevalence of antibiotic use was 24.5% (*n* = 243). As shown in Figure 1, sepsis was the most common reason for administering antibiotics (22.6%), followed by pneumonia (13.6%), surgical causes (11.5%) and upper or mild respiratory infections (9.5%). Information about the classes of antibiotics used was available for 153 (63.0%) patients. Among these, 86 (56.2%) received cephalosporines, 33 (21.6%) aminoglycosides, 33 (21.6%) penicillins, 31 (20.3%) antistaphylococcal drugs, 17 (11.1%) antianaerobic drugs, 14 (9.2%) macrolides, 11 (7.2%) carbapenems, and 1 (0.7%) fluoroquinolones. The percentage distribution of single and combination antibiotic therapies is illustrated in Figure 2. The rate of antibiotic prescriptions remained stable during the whole study period with an average decrease of –0.6% (95% CI – 2.6%, 1.3%) from April 2020 to October 2020 (Figure 3).

**Figure 1.**
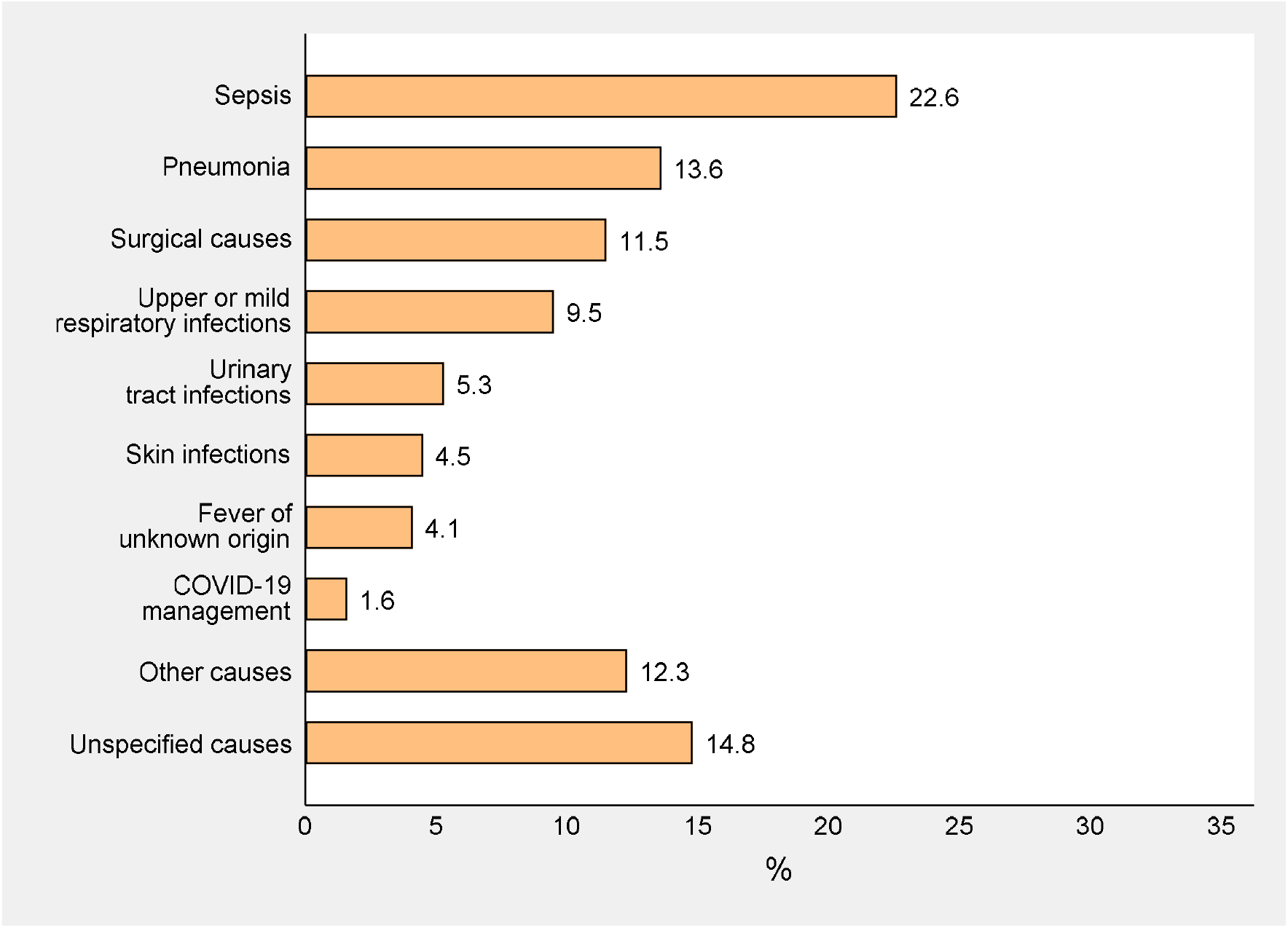
Reasons for antibiotic use (*n* = 243).

**Figure 2.**
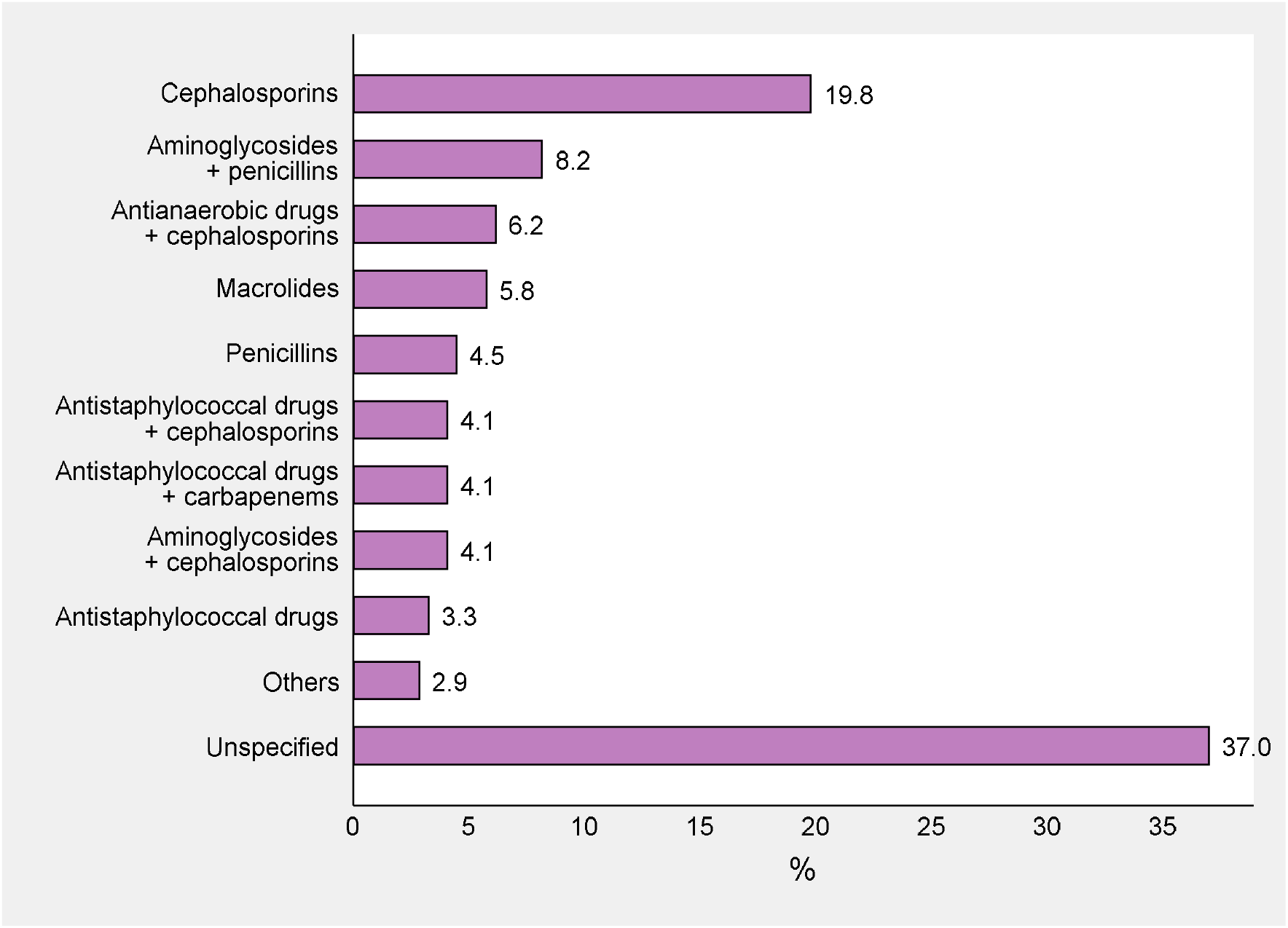
Classes of antibiotics administered to the patients, alone or in combination (*n* = 243). *Notes:* cephalosporines include cefalexine, cefalotin, cefepime, cefotaxime, ceftazidime and ceftriaxone; aminoglycosides include amikacin and gentamicin; penicillins include amoxicillin and ampicillin; antianaerobic drugs include metronidazole; macrolides include azithromycin and clarithromycin; antistaphylococcal drugs include clindamycin, trimethoprim and vancomycin; carbapenems includemeropenem; others include various combinations of these antibiotics as well as fluoroquinolones (ciprofloxacin).

**Figure 3.**
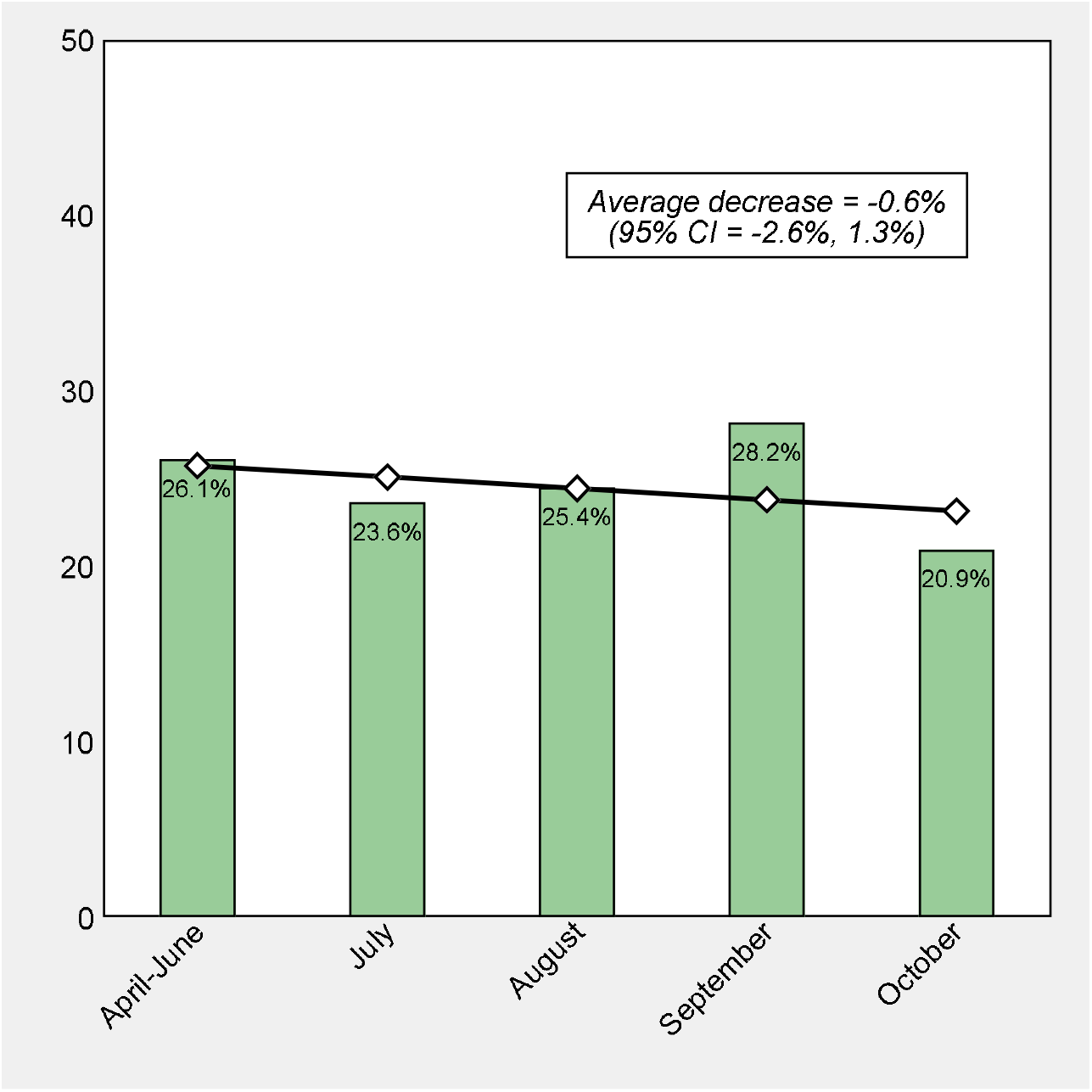
Prevalence of antibiotic use between April and October 2020. *Notes:* Linear trend was assessed using a linear regression model with variance-weighted least squares.

On multivariable analysis (Table 2), MIS-C with cardiac involvement (OR = 45.48), MIS-C with no cardiac involvement (OR = 10.35), provision of intensive care (OR = 9.60), need for hospitalization (OR = 6.87), pneumonia and/or ARDS detected through chest X-rays (OR = 4.40), administration of systemic corticosteroids (OR = 4.39), oxygen support, mechanical ventilation or CPAP (OR = 2.21), pyrexia (OR = 1.84), and female sex (OR = 1.50) were independently associated with increased use of antibiotics. On the contrary, lower respiratory tract infections not suggestive of pneumonia/ARDS and not requiring respiratory support (OR = 0.34) were independently associated with decreased use of antibiotics. We also found large and significant variations in antibiotic use across the hospitals.

**Table 2.** Multivariable logistic regression analysis of antibiotics use (*n* = 990).

| Characteristic | Odds ratio | P-value | 95% confidence interval |  |
| --- | --- | --- | --- | --- |
|  |  |  | Lower bound | Upper bound |
| Sex |  |  |  |  |
| Male | Ref. |  |  |  |
| Female | 1.50 | 0.040 | 1.02 | 2.21 |
| Age group |  |  |  |  |
| 0 y | Ref. |  |  |  |
| 1–2 y | 0.63 | 0.118 | 0.35 | 1.13 |
| 3–5 y | 0.82 | 0.556 | 0.43 | 1.58 |
| 6–11 y | 1.13 | 0.670 | 0.65 | 1.98 |
| 12–17 y | 0.92 | 0.821 | 0.47 | 1.82 |
| Immunosuppressants, immunodeficiency or chemo |  |  |  |  |
| No | Ref. |  |  |  |
| Yes | 1.65 | 0.451 | 0.45 | 6.05 |
| Hospitalization |  |  |  |  |
| No | Ref. |  |  |  |
| Yes, without intensive care | 6.87 | <0.001 | 4.34 | 10.89 |
| Yes, with intensive care | 9.60 | <0.001 | 2.77 | 33.27 |
| Pyrexia ($\geq 38.0/\geq 100.4$ °C/°F) | | | | |
| No | Ref. |  |  |  |
| Yes | 1.84 | 0.011 | 1.15 | 2.96 |
| Upper respiratory tract infection |  |  |  |  |
| No | Ref. |  |  |  |
| Yes | 1.08 | 0.730 | 0.71 | 1.65 |
| Diarrhea and/or vomiting |  |  |  |  |
| No | Ref. |  |  |  |
| Yes | 1.05 | 0.822 | 0.67 | 1.64 |
| Lower respiratory tract infection |  |  |  |  |
| No | Ref. |  |  |  |
| Yes | 0.34 | 0.007 | 0.16 | 0.74 |
| Headache |  |  |  |  |
| No | Ref. |  |  |  |
| Yes | 0.88 | 0.746 | 0.42 | 1.87 |
| Chest X-ray abnormalities (pneumonia and/or ARDS) |  |  |  |  |
| No | Ref. |  |  |  |
| Yes | 4.40 | <0.001 | 1.99 | 9.71 |
| Oxygen support, mechanical ventilation and/or CPAP |  |  |  |  |
| No | Ref. |  |  |  |
| Yes | 2.21 | 0.050 | 1.002 | 4.88 |
| Administration of systemic corticosteroids |  |  |  |  |
| No | Ref. |  |  |  |
| Yes | 4.39 | <0.001 | 2.01 | 9.58 |
| MIS-C diagnosis |  |  |  |  |
| No | Ref. |  |  |  |
| Yes, w/o cardiac involvement | 10.35 | 0.050 | 1.005 | >100 |
| Yes, w/ cardiac involvement | 45.48 | 0.011 | 2.44 | >100 |
| Center |  |  |  |  |
| Hospital San Bartolomé (PE) | Ref. |  |  |  |
| Hospital Nacional de Niños (CR) | 0.58 | 0.034 | 0.34 | 0.96 |
| Hospital Isidoro Iriarte (AR) | 0.25 | <0.001 | 0.14 | 0.46 |
| Hospital Edgardo Rebagliati Martins (PE) | 0.04 | 0.014 | 0.00 | 0.53 |
| Hospital Pablo Tobón Uribe (CO) | 0.05 | <0.001 | 0.01 | 0.25 |
| Clínica Jasmédica (PE) | 1.48 | 0.466 | 0.51 | 4.28 |
| Hospital General Regional 200 Tecámac (MX) | 12.24 | 0.045 | 1.06 | >100 |

## DISCUSSION

In our study, the prevalence of antibiotic prescribing in children with COVID-19 and MIS-C was 24.5%. We found significant variations in classes of antibiotics used and even large differences across the hospitals. The rate of antibiotic prescriptions was significantly higher in children with MIS-C, those requiring respiratory support, those with radiologic evidence of pneumonia/ARDS and those with fever. Interestingly, younger children and those with symptoms suggestive of lower respiratory tract infections without radiologic evidence of pneumonia/ARDS and not requiring respiratory support were less frequently prescribed with antibiotics. Importantly, also the only need for admission to the hospital was associated with a higher rate of antibiotic prescription. To our knowledge, this is the first multinational study assessing the use of antibiotics in children with COVID-19 and MIS-C, therefore pediatric studies to compare our findings are not available.

Velasco-Arnaiz et al (11) are the only authors that evaluated antibiotic use in a pediatric referral center before and during the pandemic. The use of azithromycin, initially considered as first-line therapy in severe COVID-19 patients in combination with hydroxychloroquine, increased, particularly in PICU setting. The use of ceftriaxone and teicoplanin, doubled in the PICU in April 2020 compared with April 2019. In non-PICU patients, piperacillin-tazobactam and ciprofloxacin use increased. Other antibiotics for community-acquired infections were prescribed less than in the same period in 2019, and cefazolin use decreased due to the dramatic drop in the number of surgeries. Also in our cohort, cephalosporins were frequently prescribed, while, interestingly, macrolides represented only 9.2% of all prescriptions. This is probably because the peak of pediatric cases in Latin America was registered when the concept of utility of azythromicine in COVID-19 was weaker.

Confirmed or suspected sepsis was the main reason for antibiotic prescription. This was an expected finding, since the pathogenesis (13) and clinical presentation of MIS-C overlap with those of sepsis, and there is general consensus for starting broad-spectrum antibiotics in these children (9). However, MIS-C children represented only 7.0% of the entire cohort, while 24.5% of children received antibiotics. These data suggest a potential overuse of empirical antibiotics in COVID-19 children. Considering that COVID-19 is often a milder disease in children compared with adults (14), the pediatric community is expected to empirically use antibiotics less frequently. However, the rate of prescriptions we detected is not widely different from those reported in adult studies. In fact, in our study, the need for hospital admission was independently associated with a higher probability of receiving antibiotic (OR 6.87, 95% CI 4.34-10.89). Addressing adult studies, Seatone et al reported that 38.3% of COVID-19 patients were prescribed antibiotics. Antibiotic prevalence was 45.0%, and 73.9% were prescribed for suspected respiratory tract infection. Amoxicillin, doxycycline and co-amoxiclav accounted for over half of all antibiotics in non-critical care wards, and meropenem, piperacillin-tazobactam and co-amoxiclav accounted for approximately half prescribed in critical care (15).

Although there are no data on bacterial co-infections in children with COVID-19 that may inform better policies of pediatric antimicrobial stewardships during the pandemic, even in adults, where COVID-19 is having a much more severe impact, the burden of bacterial co-infections seems to be relatively low in most published studies (16-23). Buehrle et al found bacterial infections in 31% (5/16) of COVID-19 patients, while antibiotics were administered to 56% (9/16) of patients during hospitalization, but 100% (9/9) of patients requiring ICU care (16). In Spain, Garcia-Vidal et al (17) found that 31/989 (3%) COVID-19 adults presented with community-acquired co-infections, mainly *Streptococcus pneumoniae* and *Staphylococcus aureus* pneumonia. Hospital-acquired infection was diagnosed in 43/989 patients (4%), with 25/44 (57%) occurring in critical care (mainly *Pseudomonas aeruginosa, Escherichia coli*, Klebsiella spp., and *Staphylococcus aureus*). Coagulase-negative staphylococci were the most common organisms causing documented bloodstream infection (7/16; 44%). Low observed rates of bacterial and fungal infection in COVID-19 patients have also been reported from the UK, where Hughes identified bacterial infection in 51/ 836 COVID-19 patients (6%) (18, 19). A review of eighteen full texts showed that 62/806 (8%) COVID-19 patients experienced bacterial/fungal co-infection during hospital admission, while on secondary analysis, 1450/2010 (72%) of patients were found to have received antimicrobial therapy (20). One Italian study even saw a reduction in *Clostridioides difficile* infections in hospitalized patients (21). In a rapid review, Fattorini et al found that only 1.3% of 522 COVID-19 patients in intensive care units, and apparently no COVID19 patients in other units, developed a healthcare-associated super-infection with antimicrobial-resistant bacteria (22, 23).

In our study, having signs or symptoms suggestive of lower respiratory tract infections, without radiologic evidence of pneumonia/ARDS, was associated with a lower probability of receiving antibiotics. This finding may be explained by the fact that in pediatrics such presentations are usually suggestive of a clinical diagnosis of bronchiolitis, wheezing or asthma, conditions that do not require routine antibiotic administration.

Our study clearly shows a high variability of reasons for antibiotic prescriptions and regimens chosen, as well as a significant variability among different centers. These findings highlight the uncertainties that physicians daily face in the management of COVID-19 patients. While the World Health Organization currently recommends against the prescribing of antimicrobials in mild to moderate COVID-19 cases without clear indication of bacterial infection (24), the difficulty in differentiating COVID-19 from bacterial infections on initial presentation challenges clinicians and antimicrobial stewardship practices (25). Almost after one year of the pandemic, there is no evidence to support decision-making on bacterial infection and antimicrobial stewardship in the context of COVID-19 (19), particularly in children. This uncertainty is likely to drive unnecessary antimicrobial prescribing in COVID-19 children who are unlikely, according to adult evidences, to benefit from empiric antibiotic prescriptions. This scenario will potentially increase the selection of drug resistant infections (26) and will make patients more vulnerable to bacterial infections, even during future viral pandemics that may favor bacterial co-infections from drug resistant bugs (27).

Our study has some limitations to address. We did not collect bacteria isolation and antibiotic sensitivities throughout the pandemic in the participating centers. Blood results, including inflammatory markers, were not collected. In addition, an independent expert did not assess the appropriateness of antibiotic prescription, nor the length of administration. Last, the reason for starting antibiotic was subjectively based on the evaluating clinician. The main reason for this approach was that Latin American clinicians are still struggling in the front-line, with hospitals having limited human resources to dedicate extra time for clinical research. Despite these limitations, this study provides the largest overview of antibiotic use in children with COVID-19 and MIS-C to date.

In conclusion, our study showed a relatively high rate of antibiotic prescriptions in children with COVID-19 and in particular in those with severe disease or MIS-C. Importantly, we found a significant variation in reasons for prescriptions of antibiotics and type of chosen therapies, as well in hospital practices, highlighting current uncertainties and lack of guidelines for the recognition of bacterial infections in COVID-19 children. Prospective studies are urgently needed to provide better evidence on the recognition and management of bacterial infections in COVID-19 children, as well as to develop dedicated antimicrobial stewardship programs.

## Data Availability

available upon request

## ACKNOWLEDGMENTS

We are grateful to all collaborators that helped the development of the DOMINGO study group.

## *Declarations

### Ethics approval

Mexico: COMINVETICA-30072020-CEI0100120160207; Colombia: PE-CEI-FT-06; Peru: No. 42-IETSI-ESSALUD-2020; Costa Rica: CEC-HNN-243-2020

### Consent to participate

approved

### Consent for publication

approved

## Availability of data and material

available upon request

## Authors’ contributions

DB conceptualized the study. JL performed statistical analyses. All authors contributed with data collection, writing of the draft of the manuscript-All authors read and approved the final version of the manuscript.

## List of Abbreviation

COVID-19: coronavirus disease 2019
SARS-CoV-2: severe acute respiratory syndrome coronavirus-2
MIS-C: multisystem inflammatory syndrome in children

